# Intentions to participate in cervical and colorectal cancer screening during the COVID-19 pandemic: a mixed-methods study

**DOI:** 10.1101/2021.07.20.21260558

**Authors:** Rebecca Wilson, Harriet Quinn-Scoggins, Yvonne Moriarty, Jacqueline Hughes, Mark Goddard, Rebecca Cannings-John, Victoria Whitelock, Katriina L Whitaker, Detelina Grozeva, Julia Townson, Kirstie Osborne, Stephanie Smits, Michael Robling, Julie Hepburn, Graham Moore, Ardiana Gjini, Kate Brain, Jo Waller

## Abstract

Worldwide, cancer screening faced significant disruption in 2020 due to the COVID-19 pandemic. If this has led to changes in public attitudes towards screening and reduced intention to participate, there is a risk of long-term adverse impact on cancer outcomes. In this study, we examined previous participation and future intentions to take part in cervical and colorectal cancer (CRC) screening following the first national lockdown in the UK.

Overall, 7543 adults were recruited to a cross-sectional online survey in August-September 2020. Logistic regression analyses were used to identify correlates of strong screening intentions among 2,319 participants eligible for cervical screening and 2,502 eligible for home-based CRC screening. Qualitative interviews were conducted with a sub-sample of 30 participants. Verbatim transcripts were analysed thematically.

Of those eligible, 74% of survey participants intended to attend cervical screening and 84% intended to complete home-based CRC screening when next invited. Thirty percent and 19% of the cervical and CRC samples respectively said they were less likely to attend a cancer screening appointment now than before the pandemic. Previous non-participation was the strongest predictor of low intentions for cervical (aOR 26.31, 95% CI: 17.61-39.30) and CRC (aOR 67.68, 95% CI: 33.91-135.06) screening. Interview participants expressed concerns about visiting healthcare settings but were keen to participate when screening programmes resumed.

Intentions to participate in future screening were high and strongly associated with previous engagement in both programmes. As screening services recover, it will be important to monitor participation and to ensure people feel safe to attend.

## Introduction

Screening programmes for cervical and colorectal cancer (CRC) are part of the mainstay of cancer control in many countries (1). The global Coronavirus pandemic saw unprecedented disruption to cancer screening in 2020, with national lockdowns and prioritisation of COVID-19 services causing many screening programmes to be paused.

In the UK, the three national cancer screening programmes were effectively paused between March and June 2020 when national lockdown restrictions were in place (2). It is estimated that around 3 million fewer people than normal in the UK had cancer screening between March and September 2020 (3). Disruption to cervical screening is predicted to cause over 600 excess cervical cancer cases in England (4) and changes across the care pathway for CRC led to an estimated 3,500 fewer people beginning treatment between April and October 2020 (5). Modelling work in the Netherlands, Canada and Australia estimates an increase of 0.2 to 0.5% in CRC mortality if screening disruption continues and highlights the need for coverage to be rapidly restored to pre-2020 levels (6). It is essential that cancer screening continues to be seen as important and that public confidence in the safety of healthcare settings remains high, to avoid falling uptake and increased cancer burden.

Little is known about the impact of the pandemic on attitudes towards cancer screening or intentions to take part. Before the pandemic, screening intentions in the UK have been consistently high (and higher than actual uptake). In a 2016 population-based survey of screening-eligible British women, 88% said they would attend when next invited (7). In a study combining four population-based surveys of 60–70 year-olds in England carried out in 2014-16, 79-84% of participants reported that they would *definitely*/*probably* complete CRC screening in the future (8). In one of these surveys, 64% of participants *definitely* intended to complete their next CRC screening kit (9). Intentions were higher in a primary care based survey of people aged 45-59 in England who were not yet eligible for CRC screening: 74% reported that they would *definitely* complete the kit (10).

Barriers vary between screening programmes, partly because cervical screening involves attending an appointment whereas CRC screening kits are posted for self-completion and involve stool sampling. Barriers to cervical and CRC screening and socio-demographic inequalities in uptake are well-established (7,11–14). The pandemic may have exacerbated existing practical barriers such as difficulty booking an appointment, and raised new emotional barriers reflecting concerns about COVID-19 infection risk and burdening the health service. In addition, it may have widened existing social inequalities, making screening harder for those disproportionately affected by the pandemic.

We present analyses from the population-based COVID-19 Cancer Attitudes and Behaviour Study (CABS) (15) measuring intentions to take part in cervical and CRC screening following the first UK national lockdown. Understanding cancer screening barriers and intentions during the pandemic is an essential first step towards mitigating potential long-term adverse effects on screening participation.

## Methods

We carried out a mixed-methods study including a cross-sectional population-based online survey and qualitative interviews with a sub-sample of survey participants. The methods are described in detail elsewhere (15, 16). The survey was carried out in August-September 2020 and the interviews in September-November 2020.

### Participants

The online survey participants were English-speaking adults (aged 18+) living in the UK and recruited via Cancer Research UK’s online panel provider, the HealthWise Wales database and social media. For this analysis, we used data from two sub-samples of survey respondents: women and people with a cervix aged 25-64 years (eligible for cervical screening) and people aged 60-74 years in England, Wales and Northern Ireland and age 50-74 in Scotland (eligible for CRC screening). Information on breast screening was only collected from participants recruited via HealthWise Wales, therefore analyses of these data are not presented here.

### Quantitative methods and analysis

Outcome measures were intention to take part in cervical and CRC screening when next invited (see (17) for exact wording). Binary variables indicating strong intention to take part were created (Yes, definitely vs. Yes, probably/No, probably not/No, definitely not/Don’t know). Responding ‘Yes, definitely’ has been shown to have a strong association with actual uptake (18).

Potential explanatory variables were sex (for CRC screening), age, ethnicity, marital status, educational attainment, smoking status, UK country of residence, disability status, pre-existing health conditions and personal or family/friends’ history of cancer. Participants were asked if they had experienced any of 13 barriers to cervical screening and 10 barriers to CRC screening (yes/no) derived from the Cancer Research UK Cancer Awareness Measure 2019 (19) with COVID-specific items developed for this study. The number of barriers endorsed was summed to create a total barriers score for cervical (range: 0-13) and CRC (range: 0-10) screening. Attitudes to attending healthcare settings and concern about delays to cancer screening and diagnosis in the context of the COVID-19 pandemic were measured using six items (20) (see Table 1). Participants who had not attended their last cervical screening were asked if this was due to coronavirus, with response options “No, not going was not related to coronavirus”, “Yes, I tried to go but wasn’t able to go due to coronavirus”, “Yes, I chose not to go due to coronavirus”.

**Table 1.**
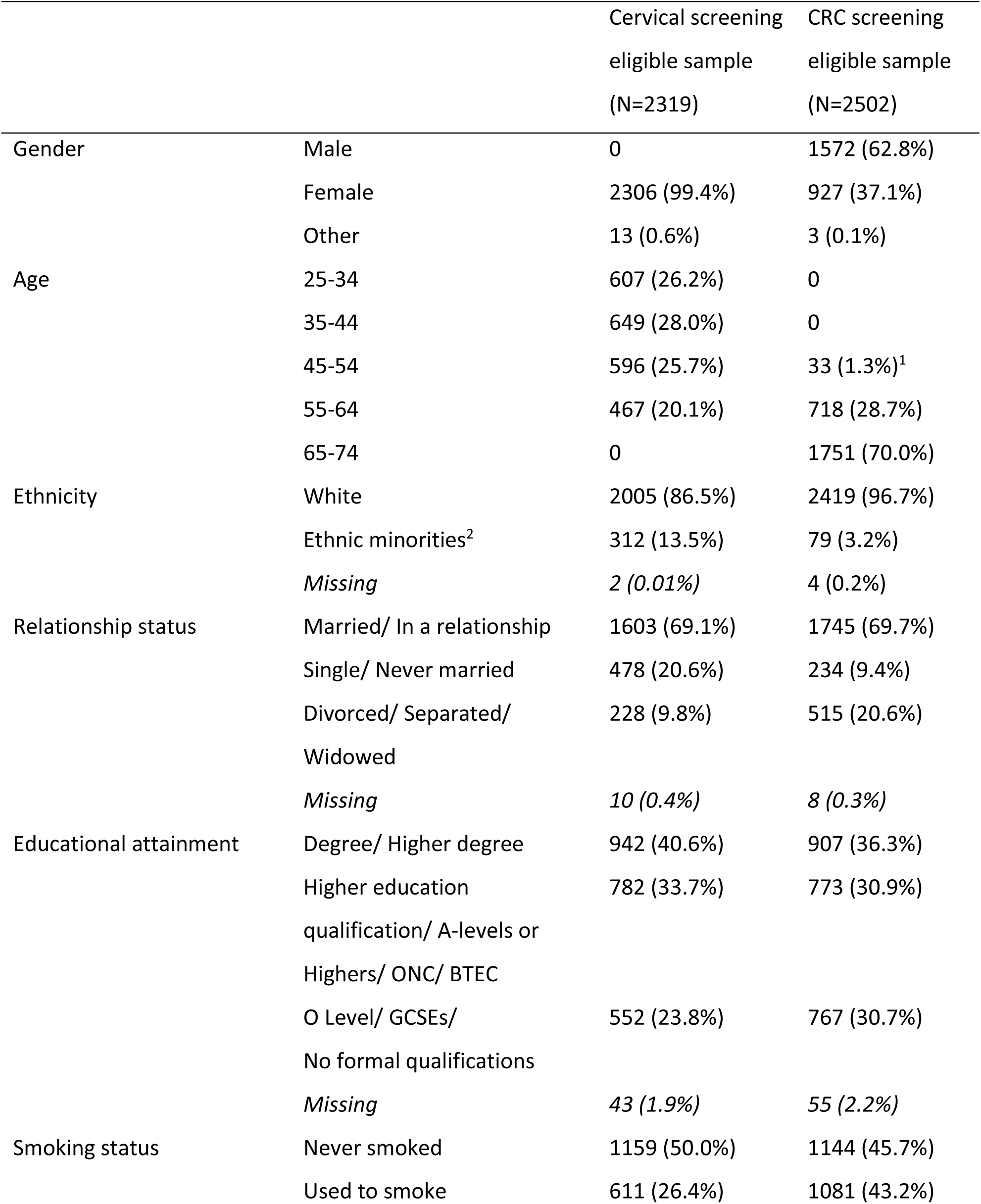
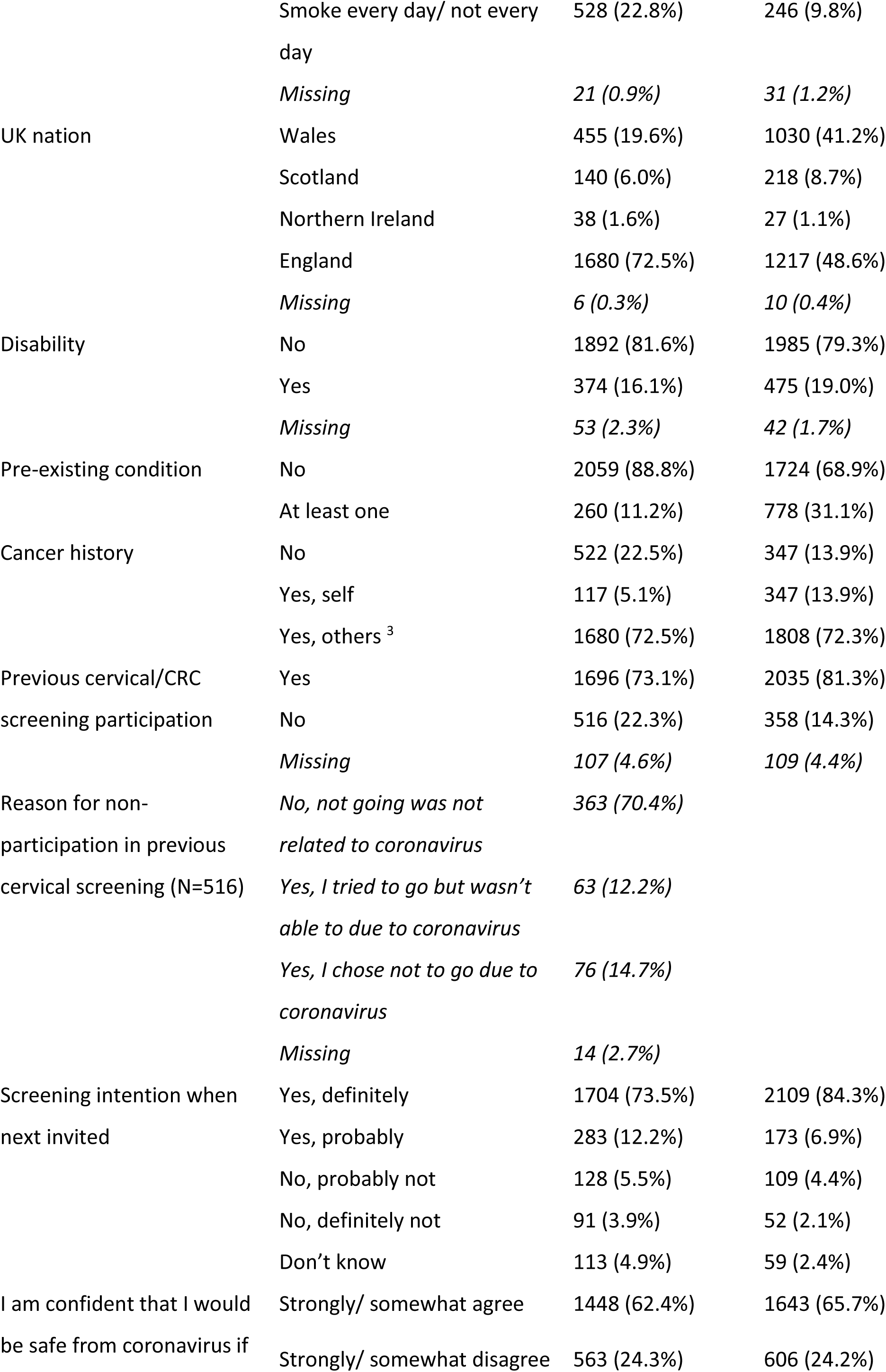
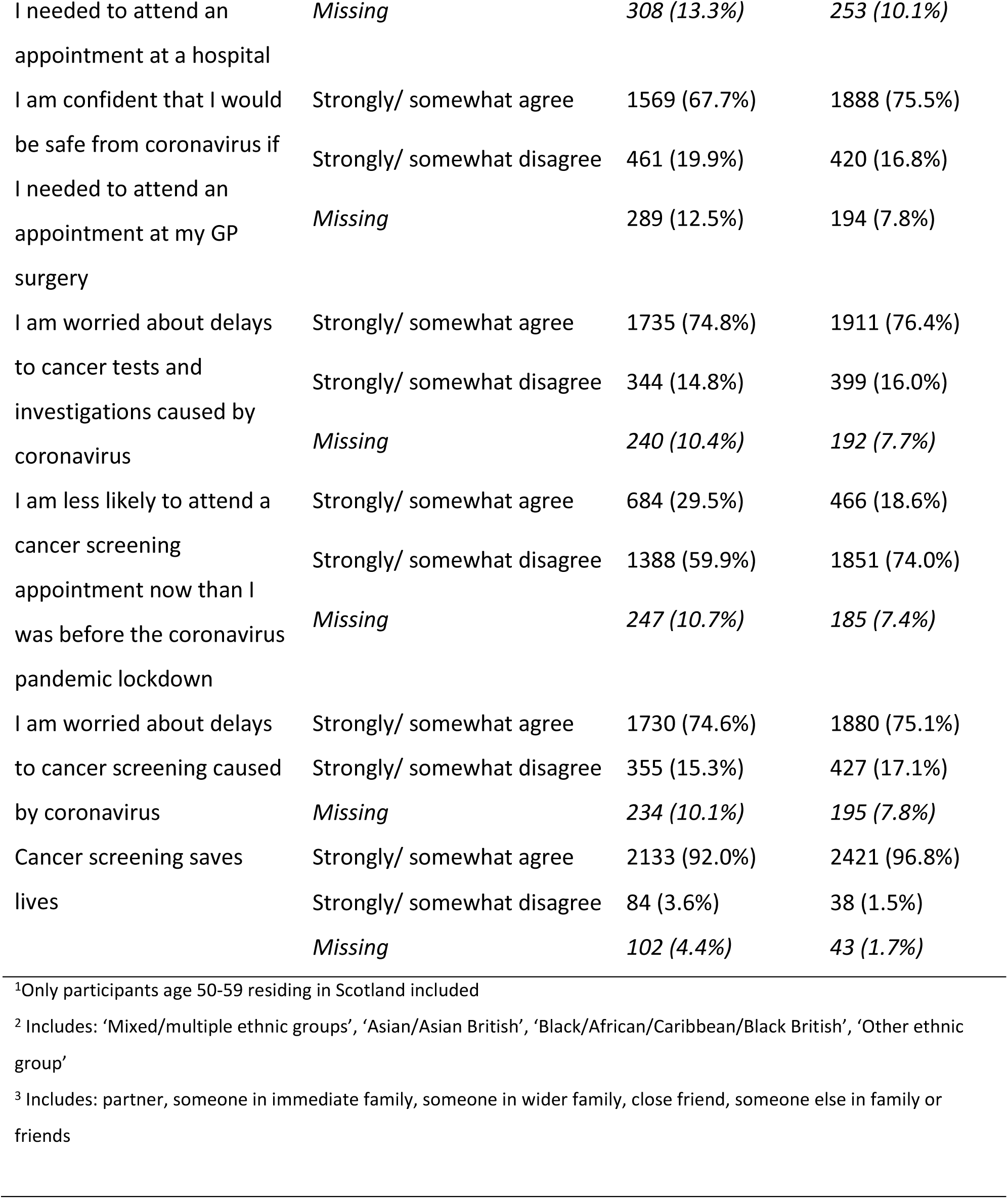
Sample characteristics for cervical screening-eligible and CRC screening-eligible participants in the UK, August-September 2020.

A pre-specified analysis plan was published on Open Science Framework (16). Descriptive statistics were used to characterise the study samples. Explanatory variables were regressed onto each binary intention outcome. Bonferroni correction was used to adjust for multiple testing and a 99.9% confidence interval (CI) was used for bivariate regression models (alpha 0.001). All explanatory variables of interest were included in the multivariable models. Sample weights were included in the bivariate and multivariable models. Analysis was performed using Stata 16 (21).

### Qualitative methods and analysis

Semi-structured telephone interviews were conducted with survey participants who had consented to be recontacted. We used purposive sampling to ensure a range of participants with respect to age, gender and symptom experience (the primary outcome in the main study). Interview participants received a £20 high-street voucher.

A topic guide was used to explore participants’ experiences of cancer symptoms and help-seeking, screening and health-related behaviours in the previous six months (Supplementary Material 1). Where participants had not been invited to or taken part in cancer screening during this period, they were asked to consider hypothetically how they might have responded if they had been invited. Interviews were transcribed verbatim and anonymised. Data were analysed thematically (22) using NVivo12 (23) with 20% double coded. The analysis presented here relates to screening-related themes only.

## Results

### Quantitative results

#### Sample characteristics

From the overall survey sample of 7543, we included 2319 participants who were eligible for cervical screening and 2502 eligible for CRC screening. Participants who preferred not to state their intentions were excluded (n=36 and n=4 for cervical and CRC screening respectively), as were those who reported that they would not be eligible for a future invitation (cervical: n=159; CRC: n=45). Participants eligible for both screening programmes (n=1003) were included in both samples. Demographic characteristics of each sample are presented in **Table 1**. Self-reported participation in screening when last invited was high for both cervical (73%) and CRC (81%) screening. Of those who had not attended their last cervical screen, 70% (363/516) said this was unrelated to COVID-19. Twelve percent (63/516) reported being unable to attend despite trying and 15% (76/516) had chosen not to attend due to COVID-19.

### COVID-related attitudes (Table 1)

As shown in Table 1, when asked how safe from coronavirus they would feel if attending an appointment at a hospital or GP surgery, over 60% of participants in both samples reported being confident they would be safe in either healthcare setting. Almost 75% in both samples reported that they were worried about delays to cancer tests and investigations, and to screening, caused by coronavirus. Thirty percent of the cervical sample and 19% of the CRC sample agreed that they were less likely to attend a cancer screening appointment now than before the pandemic. Over 90% of both samples agreed that cancer screening saves lives.

#### Cervical and CRC screening intentions

Most eligible respondents said they would ‘definitely’ participate in cervical (74%) and CRC (84%) screening when next invited (see Table 1 for a full breakdown of responses). Intention was strongly related to previous uptake: 91% of previous cervical screening attenders definitely intended to go when next invited compared with 24% of previous non-attenders. The figures were 97% and 20% for previous CRC completers and non-completers respectively. In cervical screening, intention also varied according to whether previous non-attendance was related to COVID-19. Only 14% (50/363) of women who had not attended for reasons unrelated to COVID-19 were intending to go when next invited. This figure was 36% (27/76) for those who had decided not to attend for COVID-related reasons and 70% (44/63) for those who had been unable to attend due to COVID despite having tried.

#### Barriers to cervical and CRC screening

Participants reported between 0 and 13 barriers to cervical screening (mean=0.68; median=0) and between 0 and 7 barriers to CRC screening (mean=0.23; median=0). The most frequently reported barriers to cervical screening were worry about pain (12%), a previous bad experience (9%) and embarrassment (9%), although for previous non-attenders embarrassment was the most reported barrier (23%). Worry about catching COVID-19 was not frequently endorsed (4% overall; 10% in previous non-attenders). The most common barriers to CRC screening were finding it too messy (5%), not having symptoms (4%) and embarrassment (4%). Barriers were more frequently reported by participants who had not been screened when last invited (see Supplementary Material 2).

#### Correlates of future cervical screening intention

In unadjusted analyses (see **Table 2**), low cervical screening intention was statistically significantly associated with being from an ethnic minority background (compared with being white), being single (compared with being married/in a relationship), living in England (compared with Wales), not taking part in cervical screening when last invited and endorsing more cervical screening barriers. Concerns about COVID-19 when visiting a hospital or GP surgery were associated with lower intentions to attend screening, as was being less likely to attend a cancer screening appointment now (i.e., during the pandemic) than before. Not being worried about COVID-related delays to cancer tests and screening and not agreeing that cancer screening saves lives were associated with lower intention to attend screening. In the fully adjusted model, being single, previous cervical screening non-attendance, reporting more barriers to screening and being less likely to attend a cancer screening appointment now than before the pandemic remained statistically significant predictors of low future intention.

**Table 2.**
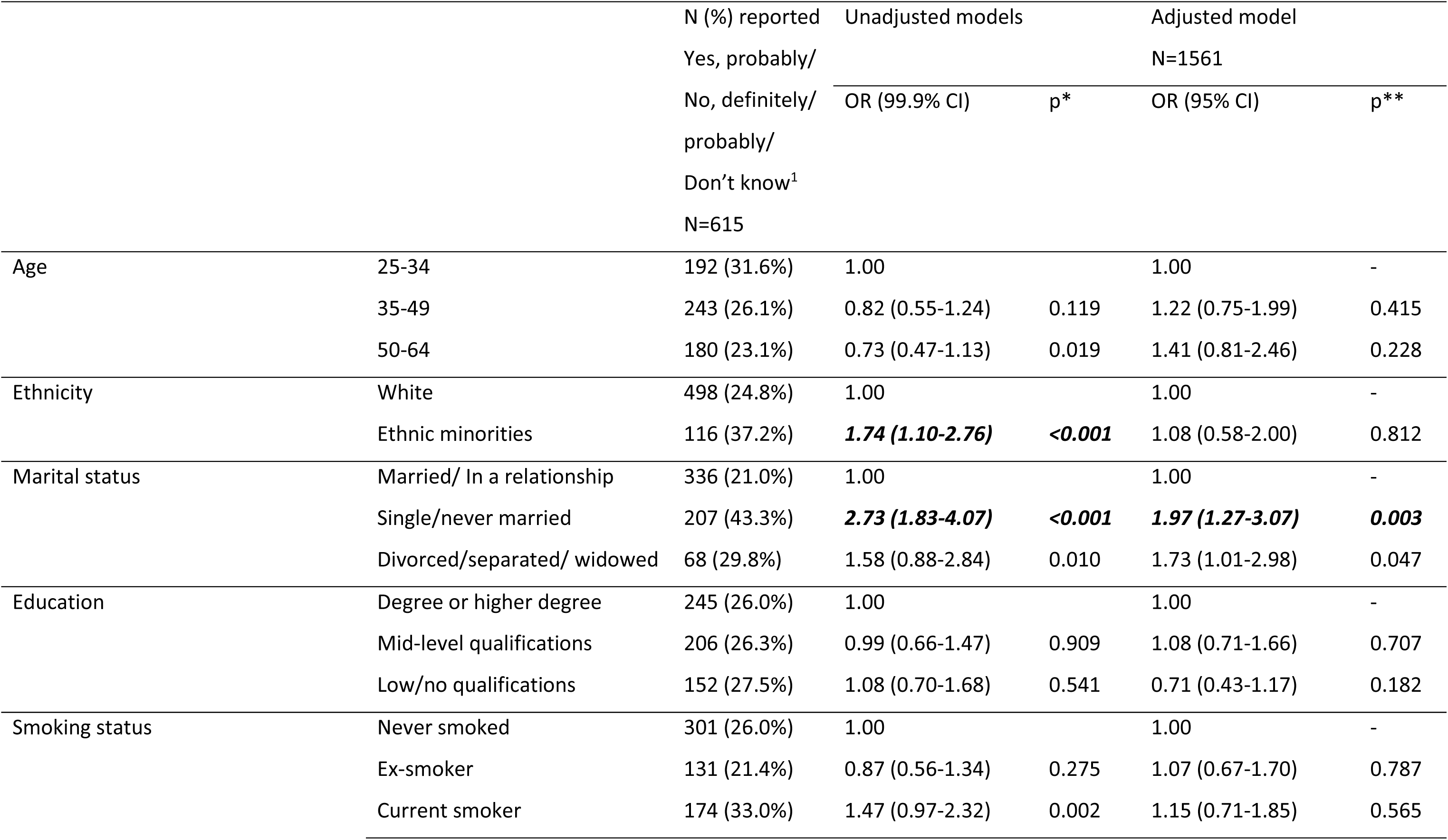
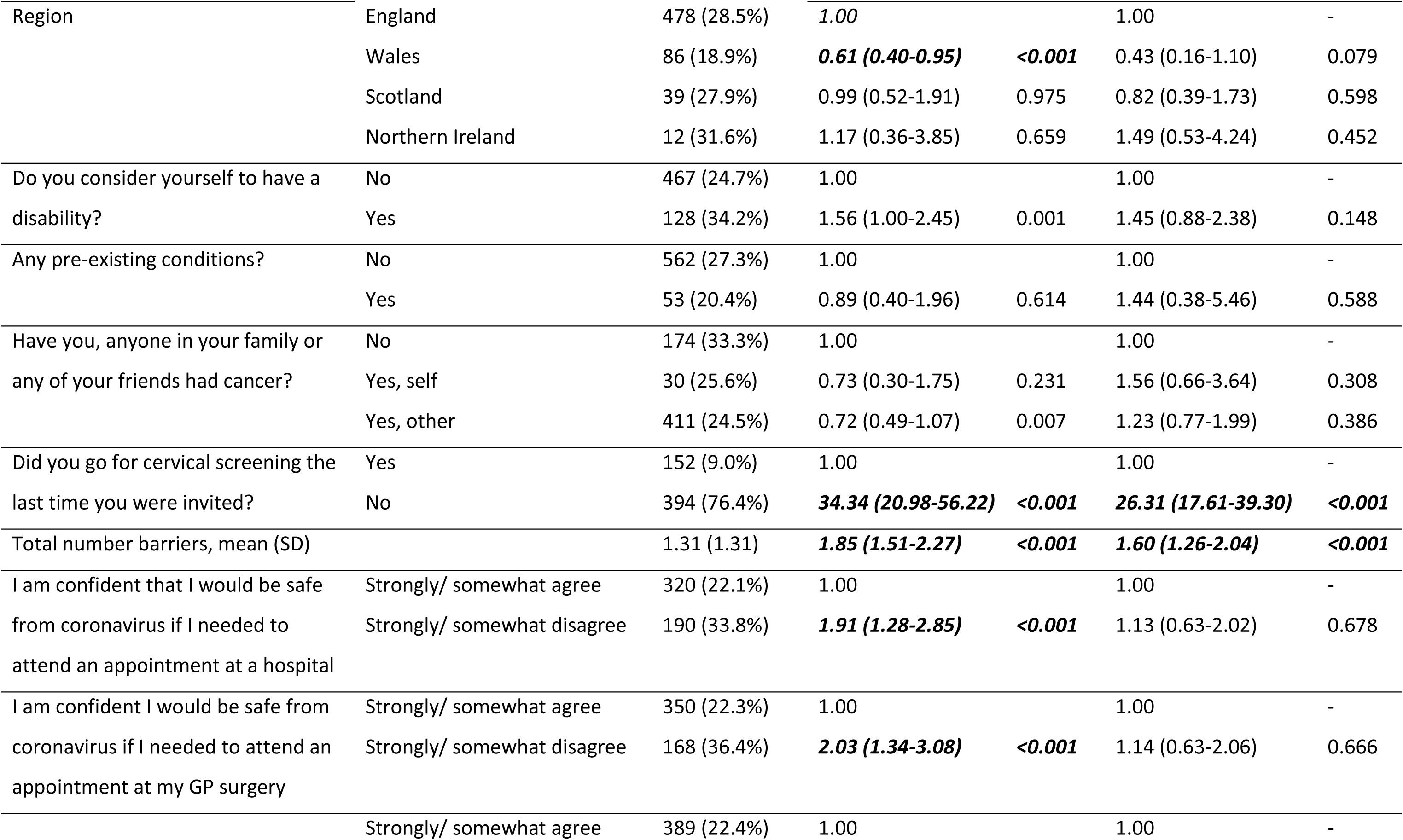
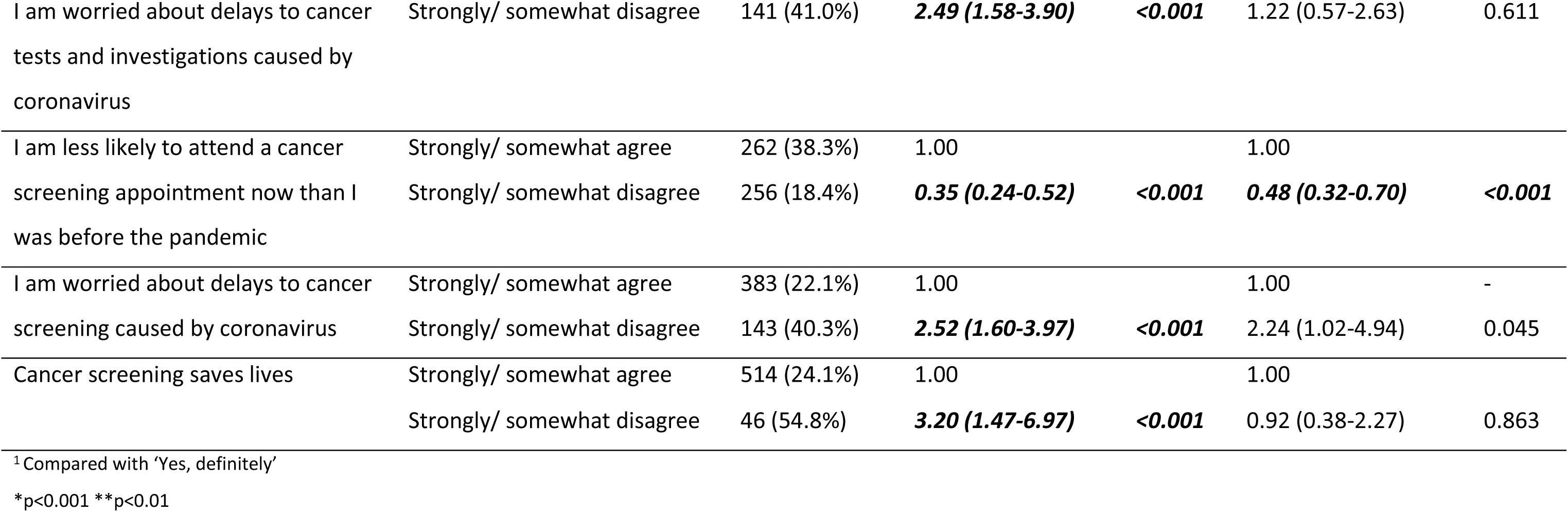
Unadjusted and adjusted logistic regression models predicting low future cervical screening intention, UK, August-September 2020 (n=2319)

#### Correlates of future CRC screening intention

In unadjusted analyses (see **Table 3**), low CRC screening intention was statistically significantly associated with being single, living in England, not having previous experience of cancer (compared with own experience or that of a friend/family), not having completed a CRC screening kit when last invited and endorsing more CRC screening barriers. Being concerned about COVID-19 when visiting the GP surgery, not being worried about COVID-related delays to cancer tests and screening and being less likely to attend a cancer screening appointment now were associated with lower intention. In the fully adjusted model, only past screening non-participation remained significantly associated with low intention to take part in future.

**Table 3.**
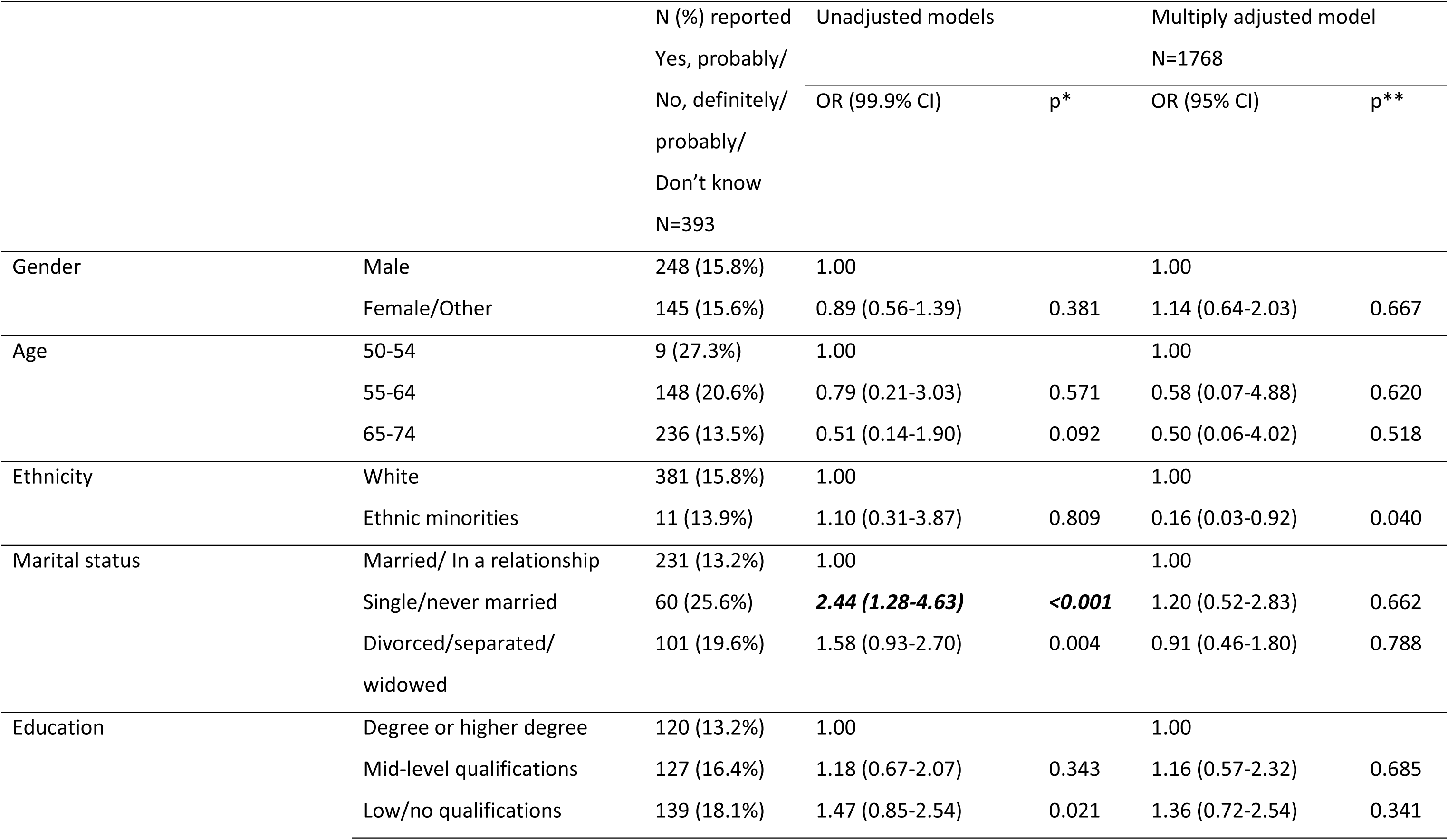
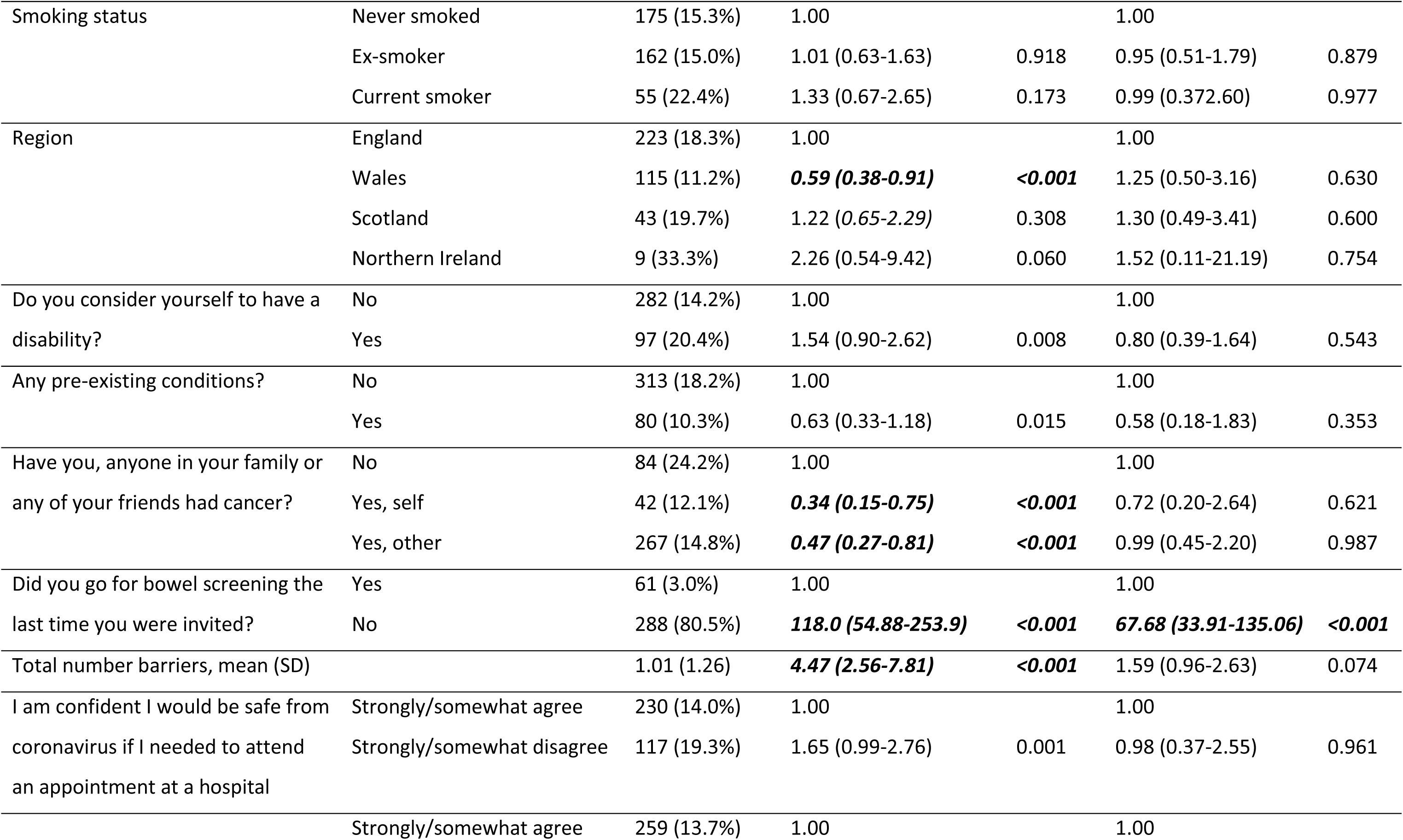
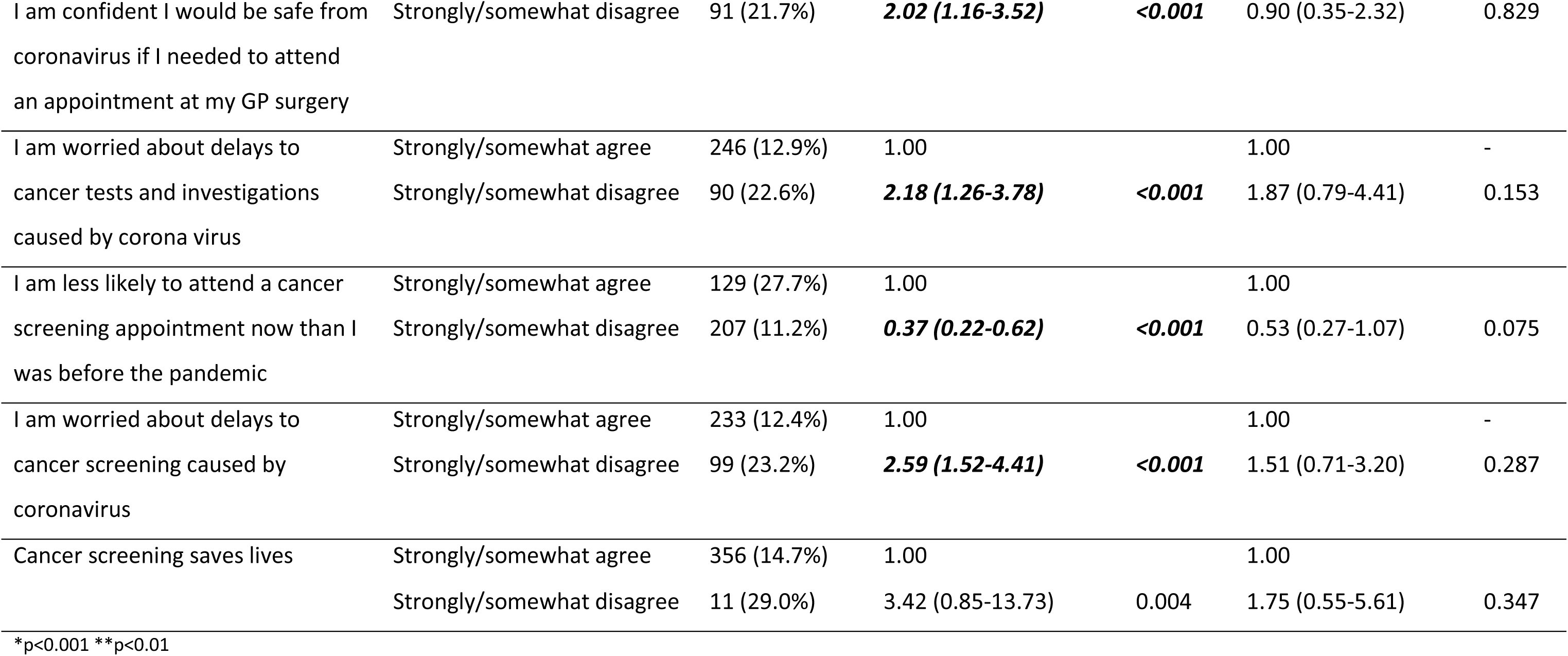
Unadjusted and adjusted logistic regression models predicting low future CRC screening intention, UK, August-September 2020 (n=2502)

### Qualitative results

#### Sample characteristics

Thirty participants were interviewed. Seventeen were male and 19 had a higher education qualification or degree. Most lived in Wales (n=25) and were from a White ethnic background (n=23). Mean age was 55 years (range: 26-76). Most were eligible for either cervical (N= 11) and/or CRC screening (N=11), with 9 not currently eligible for either. Exemplar quotes and definitions of key themes are shown in **Table 4**.

**Table 4.**
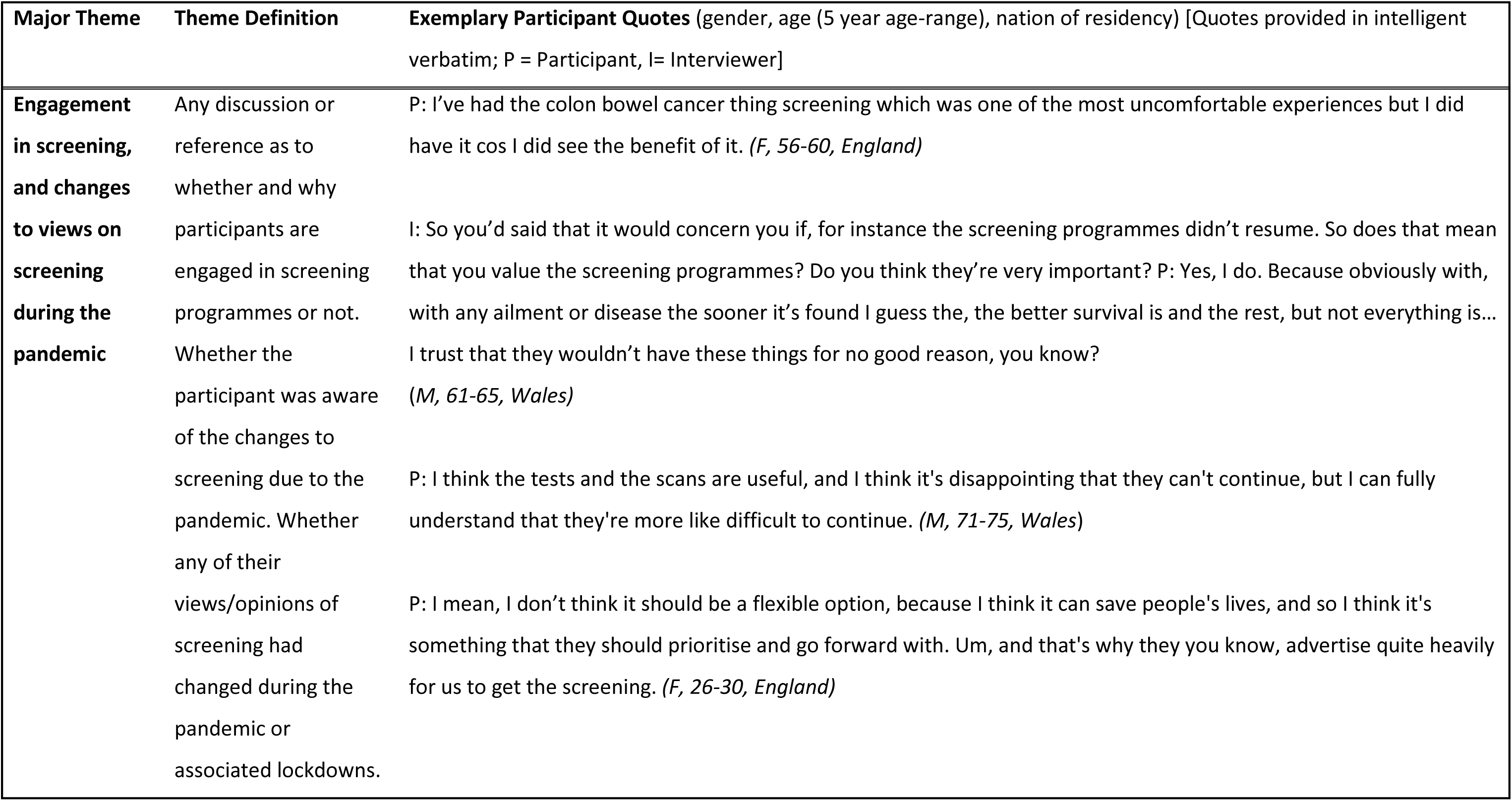
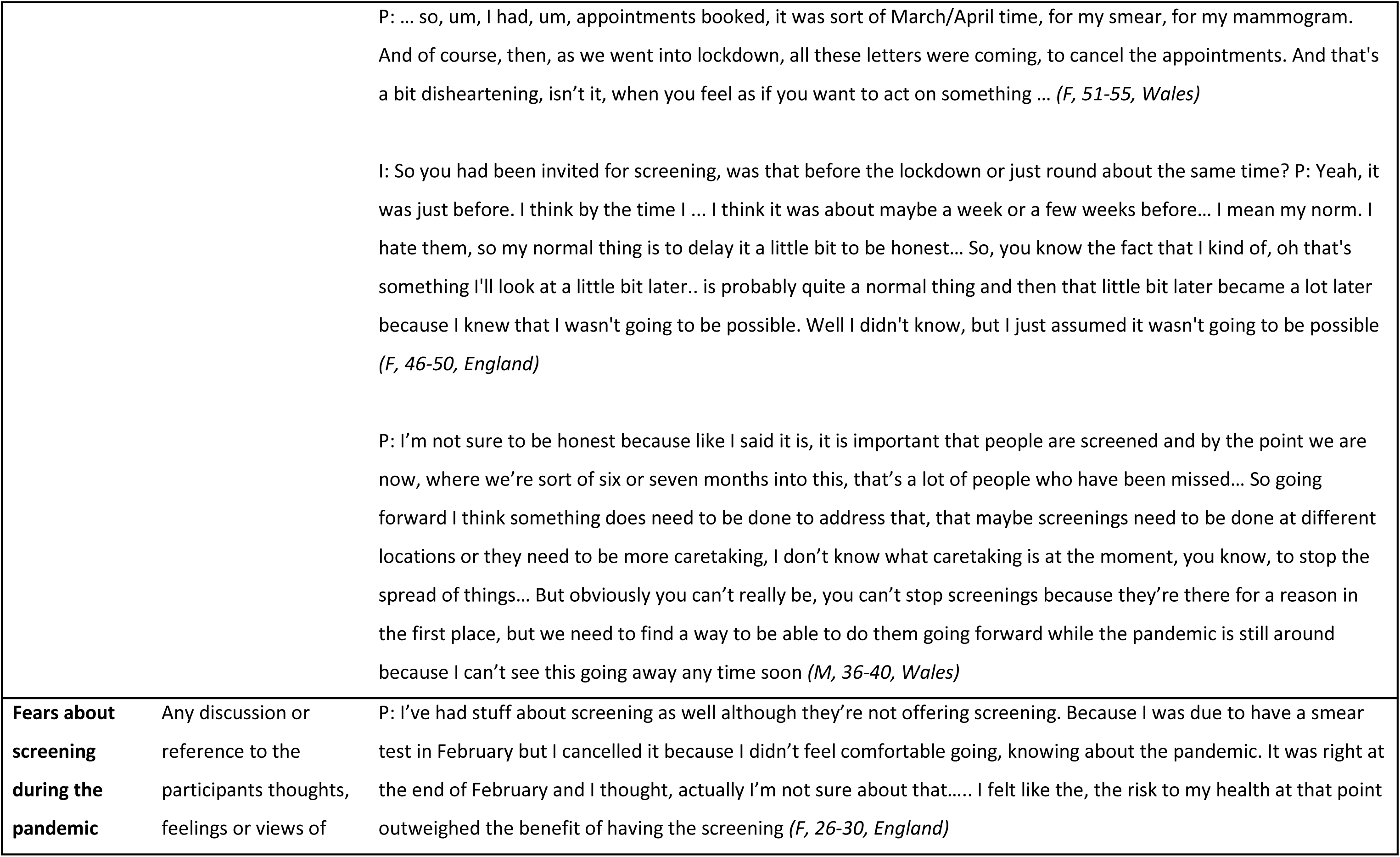
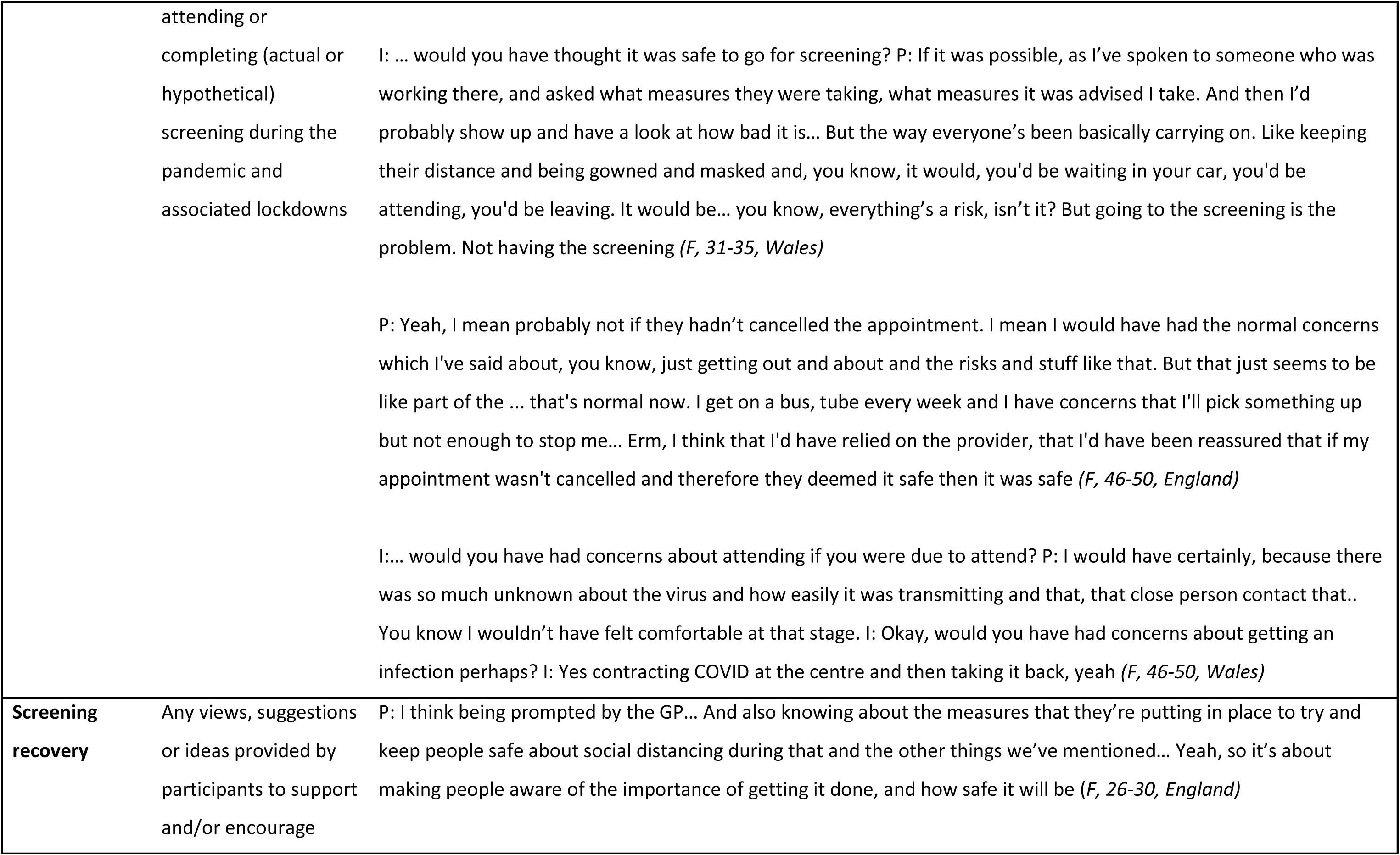
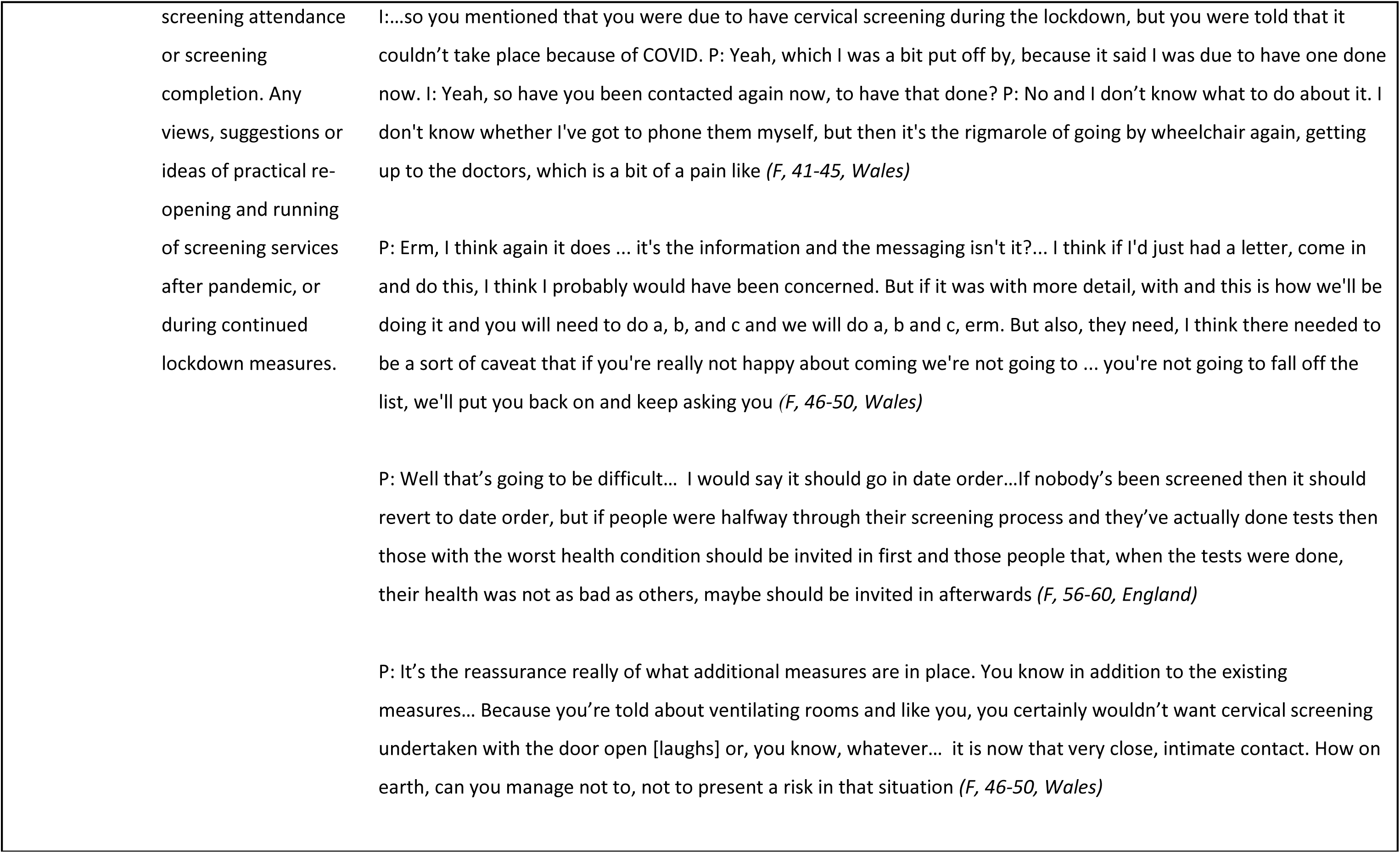
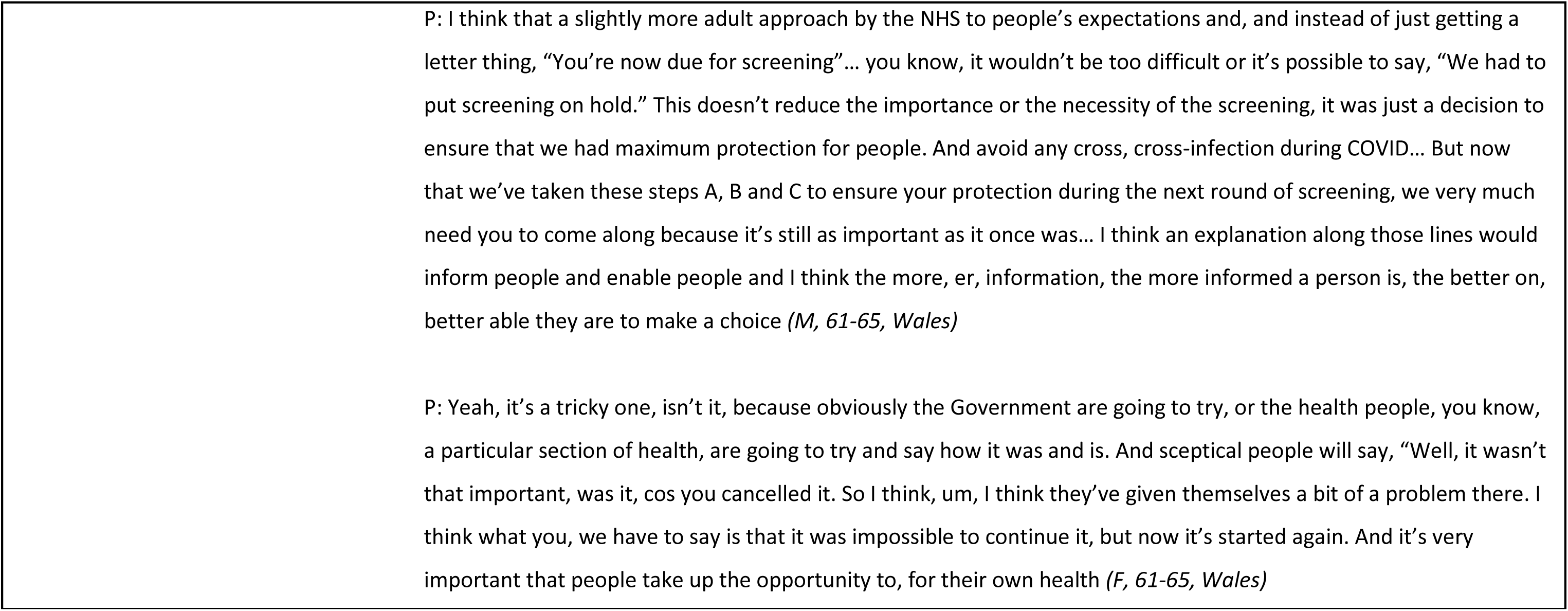
Exemplary participant quotes and theme definitions by major theme, UK, August-September 2020.

#### Views on changes to screening prior to and during the pandemic

Most eligible participants reported engaging in cancer screening prior to the pandemic. Participants were supportive of the national screening programmes and considered them important.

There was varying awareness that screening had effectively been paused. Those who were aware often understood the decision, though many expressed hope that lessons could be learned to avoid pausing screening again in the future. Participants’ understanding of the pause was frequently linked to their trust in the NHS; they trusted that doctors had made the right decision. Others questioned the need for changes as procedures and tests are often performed away from hospitals in the GP practice. These participants also expressed concerns that although COVID-19 was a priority, other conditions should not be neglected.

Participants reiterated the message that the public should be treated as adults, valuing open and honest communication about decisions on screening. They felt that this had not happened.

#### Fears about screening during the pandemic

Screening fears were related to the need to attend at a healthcare setting. Participants were not fearful of completing the CRC screening test if they had received one. Though participants mentioned some of the well-known barriers to completing the stool sample such as it being unpleasant, they expressed no specific COVID-related concerns. However, worries were heightened over cervical screening attendance (though not the procedure itself) due to its completion in a healthcare setting. This was the same for those who had been invited to screening during the pandemic and those who were asked hypothetically.

There was general fear or nervousness about attending healthcare settings. Fears were associated with the ‘unknown’ (understanding and negotiating new systems), catching COVID-19 or passing it on to others, encountering other members of the public not adhering to social distancing and using public transport. Participants described having to weigh up the risks of attendance versus not attending in making their decision.

Some of those affected voiced relief that their cervical screening appointment had been delayed or cancelled on their behalf, relieving them of the responsibility of deciding. Others expressed that to help relieve pressures for the NHS in the future (due to potential delayed diagnosis) the least they could do is to attend cervical screening when invited.

#### Screening recovery

Participants who had had screening delayed expressed confusion about whether they would be contacted to rearrange screening appointments, or whether they themselves should proactively contact services. Enthusiasm for rearranging appointments was hindered by worry that they would be bothering already stretched services. To counteract fears of COVID, participants suggested that GP practices provide information on their new infection control practices, so patients knew what to expect. When considering the backlog of those who missed screening, consensus was that this should be managed based on clinical need and risk.

## Discussion

The COVID-19 pandemic has led to widespread disruption of cancer screening. The current study is, to our knowledge, the first to show high future intentions to take part in cervical and CRC screening in the UK during the pandemic. Past non-attendance and (for cervical screening) endorsement of barriers were the strongest correlates of low future intentions to take part. For cervical screening, intention to participate was lower among women who had not attended their last screen for reasons unrelated to COVID-19. Despite overall intentions being high, a substantial minority of participants stated that they were less likely to attend a screening appointment than before the pandemic.

Our finding that 74% of women *definitely* intended to go for cervical screening (rising to 86% if the ‘yes, probably’ response was included) is comparable with Marlow et at (2017) who found 88% of women in England intended to be screened using a yes/no outcome. For CRC screening, 84% of our sample *definitely* intended to take part, which is considerably higher than the 64% reported in Dodd et al’s (2019) survey using similar methods. Despite these reassuringly high levels of intention, the finding that 30% of the cervical-eligible and 19% of the CRC-eligible samples said they were less likely to attend a screening appointment now than prior to the pandemic is concerning. This reticence is more relevant to cervical screening than home-based CRC screening and may go some way to explaining the unexpectedly lower intention for cervical than CRC screening (which is at odds with data on actual screening uptake in the two programmes; uptake rates in England were 72% for cervical screening in 2019/2020 and 64% for CRC screening in 2020 (24, 25).

Future cervical screening intentions were higher in women who reported not attending their last cervical screening for COVID-related reasons, compared with those whose reasons were not related to the pandemic. However, intentions among women who chose not to attend were lower than those who were unable to attend due to COVID-19. In addition, lower intention was associated with being less likely to attend a cancer screening appointment than before the pandemic. Directly experiencing screening disruption during the pandemic may therefore have had an impact on willingness to attend in the future. It is encouraging that screening in the UK has been much less disrupted during subsequent lockdowns but efforts may be needed to ensure those who have missed cervical screens are provided with additional support. Interview participants expressed uncertainty over the process for rebooking missed cervical screening and wished to resume screening when available.

In multivariable analyses, screening intentions were no lower in participants who were worried about visiting healthcare settings during the pandemic than those who were not worried. However, qualitative data suggested that some people did have COVID-related concerns including travelling to and attending healthcare settings, and fear of COVID-19 infection (26). As previous screening uptake was high in this sample, few barriers to screening were reported in the survey. Barriers that were endorsed mirrored those reported in pre-pandemic research including disgust, embarrassment, prior screening experiences and fear of what might be found (12,13,27,28). High levels of worry about the delays to cancer screening caused by COVID-19 were reported in the survey (though they were not associated with intentions). Recent research has shown the potentially damaging effects of the screening programme pause on cancer detection and survival (3,5,29) and participants’ concerns reflected this, perhaps due to media coverage of the impact of the pandemic on cancer services. Of note, data collection for this study (both survey and interview) took place after the first lockdown, although in the latter half of 2020 local lockdowns may have been in place for some participants. Although healthcare visits were permitted throughout lockdowns, attitudes and priorities may have been affected by local restrictions and the ‘stay at home’ message could have been interpreted as meaning that screening should wait.

### Study limitations and strengths

We recruited a large sample from across the UK and weighted data to be representative of the UK for age, gender, ethnicity and region. The data are cross-sectional, and caution must be exercised when interpreting the results. In addition, participants’ self-reported intentions might not translate into behaviour (30), although we used ‘yes, definitely’ to indicate positive intention which has demonstrated a strong association with screening behaviour (18). Intention to complete CRC screening was higher than population-level uptake, and may also reflect the high number of HealthWise Wales participants in the CRC cohort, who may be more health-motivated than the general population. (31) Sample selection bias may also explain high overall screening intentions, with limited power to detect the effects of factors expected to be associated with screening intentions such as education and ethnicity. Interview participants were sampled based on symptom experience (the primary study outcome), hence some were not eligible for screening and their responses were therefore hypothetical.

### Implications

The finding that past screening non-attendance was associated with lower future screening intention is not new, but it underlines the continued importance of interventions to reduce COVID and non-COVID screening barriers among non-participants, including women who have not attended cervical screening for COVID-related reasons. During August and September 2020 a significant minority of the population remained wary of visiting healthcare settings due to coronavirus, therefore clear public health messaging is needed to provide reassurance about the safety of attending resumed cancer screening services, prioritising known risk-groups (by disease, socioeconomic group and age) (32). It is also important to ensure there is sufficient screening and diagnostic workforce capacity, so that follow-up tests can be done in a timely manner. In addition, new adapted technologies for cervical screening have the potential to overcome a range of well-established barriers as well as reluctance to attend healthcare settings due to coronavirus risk. The pandemic provides an impetus for expedited implementation of HPV self-sampling to address cervical screening backlogs (33), potentially also increasing screening uptake.

### Conclusions

Despite concerns about attending healthcare settings during the COVID-19 pandemic, intentions to take part in cancer screening appeared to remain high during the pandemic. Efforts to restore screening participation to at least pre-pandemic levels will require clear communication with the public to address safety concerns, as well as strategies to increase screening and diagnostic workforce capacity. Ongoing evaluation is needed to assess whether high intentions are reflected in screening uptake.

## Data Availability

De-identified participant data will be made available to the scientific community with as few restrictions as feasible, whilst retaining exclusive use until the publication of major outputs. Data will be available via the corresponding author.

## Abbreviations

CABS: Cancer Attitudes and Behaviour Study
CRC: colorectal cancer
aOR: Adjusted odds ratio
GP: General Practitioner

## Additional information

Registration: ISRCTN17782018

## Acknowledgements

This study was facilitated by HealthWise Wales, the Health and Care Research Wales initiative, which is led by Cardiff University in collaboration with SAIL, Swansea University. HQS is funded by PRIME Centre Wales, which is funded by Welsh Government through Health and Care Research Wales. DECIPHer and The Centre for Trials Research receives funding from Health and Care Research Wales, and Health and Care Research Wales and Cancer Research UK respectively. SS is funded by a Health and Care Research Wales fellowship (514366). JW is funded by a Cancer Research UK Career Development Fellowship (C7492/A17219). We are grateful to Cancer Research UK’s Cancer Insights Patient Panel, Clinical Advisory Panel and GP Panel for their helpful feedback. Cancer Research UK staff members and other researchers contributed to the initiation and development of the CAM, both historically and for this project. COVID-CAM data was provided by Cancer Research UK who collected the data via Dynata’s online survey panels. External Scientific Advisory Group members who advised the statistical analysis plan include: Professor Jamie Brown (University College London), Professor Yoryos Lyratzopoulos (University College London), Dr Katie Robb (University of Glasgow), Dr Christian von Wagner (University College London) and Professor Fiona Walter (University of Cambridge).

## Author contributions

KB – Conceptualisation; Methodology; Writing – Review & Editing; Supervision; Funding acquisition

RC-J – Conceptualisation; Methodology; Validation; Formal analysis; Writing – Review & Editing; Supervision

AG – Conceptualisation; Methodology; Writing – Review & Editing

MG – Investigation; Data curation; Writing – Review & Editing

DG – Methodology; Validation; Formal analysis; Data curation; Writing – Review & Editing

JH – Methodology; Formal analysis; Writing – Review & Editing

GM – Conceptualisation; Methodology; Writing – Review & Editing

YM – Methodology; Formal analysis; Data curation; Writing – Review & Editing; Project administration;

KO – Conceptualisation; Methodology; Writing – Review & Editing MR – Conceptualisation; Methodology; Writing – Review & Editing

HQ-S – Conceptualisation; Methodology; Formal analysis; Data curation; Writing – Review & Editing; Supervision

SS - Writing – Review & Editing

JT – Conceptualisation; Methodology; Writing – Review & Editing

JW – Conceptualisation; Methodology; Writing – Review & Editing

KLW – Conceptualisation; Methodology; Writing – Review & Editing

VW – Methodology; Investigation; Data curation; Writing – Review & Editing

RW – Validation; Formal analysis; Writing – Original draft; Writing – Review & Editing

## Ethics approval

Ethical approval was granted by the School of Medicine Research Ethics Committee, Cardiff University (ref 20.68).

