## Supplementary materials for "Intentions to participate in cervical and colorectal cancer screening during the COVID-19 pandemic: a mixed-methods study"

### Supplementary Material

#### Supplementary material 1 - Interview Topic Guide

Participants will have access to study Information sheets and will have already provided informed consent. Prior to interview commencement the interviewer will re-confirm verbal consent.

The interview will be recorded, anonymised, transcribed confidentially and analysed by members of the research team.

The aim of the interview is to gain further understanding of how participants perceive symptoms, help seeking and behaviour regarding potential cancer symptoms during the COVID-19 lockdown from March 23rd 2020.

It is estimated the interview will be 45 minutes in length. Following the interview participants will be sent a £20 voucher to thank them for their time.

##### Topic Guide

**<u>Symptoms and Help seeking</u>**

- Participants' views on symptoms they may have noticed during lockdown and any medical help seeking
  *(The survey asked about symptom experience with no reference to cancer, to avoid influencing responses – the same applies in the interview)*

- For symptoms experienced, participants will be asked for more details, probes regarding timescale of help seeking, perceptions of access to primary care and what influenced their perceptions

- Participants' views about seeking medical help from a health care professional during lockdown, and what this experience was like compared to pre-lockdown consultations including any experience of remote consulting

**<u>Screening</u>**

- Participants' views on how routine cancer screening programmes were affected during lockdown (breast, bowel, cervical as applicable), their views on screening having been paused, and how has this impacted the relative importance of cancer/cancer screening in the context of pandemic concerns

- What would encourage participants to consider taking part in cancer screening when it resumes, and what may put them off

**<u>Health behaviours and Prevention</u>**

- Participants' views about any changes to their health-related behaviour (particularly smoking) and what may have influenced any changes (including perceptions of links between coronavirus outcomes and smoking)

- Participants’ views on sources of health information and health messaging during lockdown, and their perceived usefulness and credibility

Supplementary material 2 – Screening barriers

**Supplementary Table 1: Self-reported barriers to previous cervical and CRC screening participation, UK, August-September 2020**

| Thinking about the last time you were invited for cervical screening, did any of the following put you off going? | Endorsed barrier (N=2139) | Previous attenders (N=1696) | Previous non-attenders (N=516) |
|---|---|---|---|
| I was worried that cervical screening might be painful | 274 (11.8%) | 175 (10.3%) | 97 (18.8%) |
| I have had a bad experience of cervical screening in the past | 201 (8.7%) | 113 (6.7%) | 86 (16.7%) |
| I was too embarrassed to go for cervical screening | 215 (9.3%) | 93 (5.5%) | 118 (22.9%) |
| I didn’t want a man to carry out the screening test | 151 (6.5%) | 101 (6.0%) | 48 (9.3%) |
| I didn’t have any symptoms of cervical cancer | 121 (5.2%) | 72 (4.3%) | 44 (8.5%) |
| I was too frightened of what the test might find | 110 (4.7%) | 66 (3.9%) | 43 (8.3%) |
| I was worried about catching coronavirus if I went for screening | 95 (4.1%) | 40 (2.4%) | 52 (10.1%) |
| I don’t think that I am at risk of cervical cancer | 103 (4.4%) | 43 (2.5%) | 58 (11.2%) |
| I was too busy to go for cervical screening | 86 (3.7%) | 34 (2.0%) | 52 (10.1%) |
| I was too afraid of having treatment if I was found to have cancer | 74 (3.2%) | 44 (2.6%) | 29 (5.6%) |
| After thinking about the test, I decided that the risks of taking part outweigh the benefits | 64 (2.8%) | 30 (1.8%) | 29 (5.6 |
| I had other more important things to worry about than cervical screening | 60 (2.6%) | 22 (1.3%) | 36 (7.0%) |
| I had symptoms that might have been related to coronavirus | 15 (0.7%) | 5 (0.3%) | 10 (1.9%) |
| **Thinking about the last time you were invited for bowel screening, did any of the following put you off going?** | **Endorsed barrier (N=2502)** | **Previous completers (N=2035)** | **Previous non-completers (N=358)** |
| I found it too messy to complete the stool test kit | 119 (4.8%) | 27 (1.3%) | 91 (25.4%) |
| I didn’t have any symptoms of bowel cancer | 91 (3.6%) | 32 (1.6%) | 59 (16.5%) |
| I found it too embarrassing to complete the stool test kit | 88 (3.5%) | 18 (0.9%) | 69 (19.3%) |
| I found it too difficult to complete the stool test kit | 61 (2.4%) | 11 (0.5%) | 49 (13.7%) |
| I was too frightened of what the stool test might find | 59 (2.4%) | 15 (0.7%) | 44 (12.3%) |
| After thinking about the test, I decided that the risks of taking part outweigh the benefits | 35 (1.4%) | 21 (1.0%) | 14 (3.9%) |
| I had other more important things to worry about than bowel screening | 36 (1.4%) | 6 (0.3%) | 30 (8.4%) |
| I was too afraid of having treatment if I was found to have cancer | 30 (1.2%) | 12 (0.6%) | 18 (5.0%) |

| | | | |
|---|---|---|---|
| I was too busy to complete the tool test kit | 29 (1.2%) | 5 (0.3%) | 23 (6.4%) |
| I don't think that I am at risk of developing bowel cancer | 28 (1.1%) | 6 (0.3%) | 21 (5.9%) |
